# Covid-19 Social Distancing Interventions by State Mandate and their Correlation to Mortality in the United States

**DOI:** 10.1101/2020.08.26.20182758

**Authors:** Sean McCafferty, Sean Ashley

## Abstract

**Background:** Evaluate the correlation between U.S. state mandated social interventions and Covid-19 mortality using a retrospective analysis of Institute for Health Metrics and Evaluation (IHME) data.

**Methods:** Twenty-seven (27) states in the United States were selected on June 17, 2020 from IHME data which had clearly defined and dated establishment of statewide mandates for social distancing measures to include: School closures, Prohibition on mass gatherings, business closures, stay at home orders, severe travel restrictions, and closure of non-essential businesses. The state Covid-19 mortality prevalence was defined as total normalized deaths to the peak daily mortality rate. The state mortality prevalence was correlated to the total number of mandates-days from their date of establishment to the peak daily mortality date. The slope of the maximum daily mortality rate was also correlated to mandate-days.

**Results:** The standardized mortality per state to the initial peak mortality rate did not demonstrate a discernable correlation to the total mandate days (R^2^ = 0.000006, p= 0.995). The standardized peak mortality rate per state suggested a slight correlation to the total mandate days (R^2^ = 0.053,p=0.246), but was not statistically significant. There was a significant correlation between standardized mortality and state population density (R^2^ = 0.524,p=0.00002).

**Conclusions:** The analysis appears to suggest no mandate effective reduction in Covid-19 mortality nor a reduction in Covid-19 mortality rate to its defined initial peak when interpreting the mean-effect of the mandates as present in the data. A strong correlation to population density suggests human interaction frequency does affect the total mortality and maximum mortality rate.

**Trial Registration:** Public data is not patient specific and made available for public download on IHME Websites which can be used, shared, modified or built upon by non-commercial users via the Creative Commons Attribution-NonCommercial 4.0 International License. https://creativecommons.org/licenses/by-nc/4.0/

**Précis:** The mean-effects of Covid-19 mandated social interventions were found to have no affect the maximum slope of the daily mortality nor the overall mortality but did correlate to the population density.

## Background

Socially distancing policy has been theorized to effectively reduce the rate of transmission of contagious diseases.^1,2,3^ Mandating social distancing state-wide within the United States has been instituted to varying degrees during the Covid-19 pandemic. Reducing the maximum contagion transmission by mandating decreased social interactions has been theorized to reduce mortality by allowing for social/medical mobilization and proper resource allocation.^3,4,5^ Verification of this theoretical model has not been demonstrated, except analyses demonstrating marginal social distancing effectiveness in delaying the peak infection.^6^ The extensive global incidence of the Covid-19 pandemic and unique realtime record keeping presents an opportunity to evaluate the social distancing theory over a large population with considerably different social distancing measures instituted.

## Methods

The study was conducted using the Institute for Health Metrics and Evaluation (IHME) openly published data on Covid-19 infections by individual states in the United States, to include daily infections/deaths as well as onset and discontinuation dates of state mandated social interaction interventions. As this was the primary source of information used for predictive modeling and setting public policy, it was chosen for its accuracy and regular updates.^7^

The methods utilized are an extension of the recently submitted manuscript evaluating European countries in the same manner. The methods are discussed extensively in the available preprint manuscript:”Covid-19 Social Distancing Interventions by State Mandate and Their Correlation to Mortality”.^8^

The purpose was to quantify the decrease in the infection/mortality or decrease in the maximum infection/mortality rate as a result of defined state mandated social distancing measures. Mortality was chosen to define endpoint peaks and rates of change over registered infections within a state. Covid-19 registered infections are beset by inaccuracies due to: Inaccurate testing, testing frequency, asymptomatic patients, test availability, and regional variations in testing criteria. Individual states routinely record the cause and time of death. The analyses account for the state’s stage in the epidemiological infectious cycle and to extend those techniques to the states in the USA while additionally accounting for population density.

IHME data accessed on June 17, 2020 at 1900 EST was used to select all U.S. states with more than a maximum mortality rate of 10 Covid-19 deaths per day. Each of the selected states independently instituted preventive social interaction law on a given date to include one of the six (6) possible mandates.

Considered forms of social mandates:

1. Public School Closures
2. Social Gathering Restrictions
3. Stay-at-Home Orders
4. Business Closures
5. Non-Essential Business Closures
6. Severe Travel Restrictions

States which did not maintain their state-wide mandated social distancing through the end of the examination period (initial peak mortality rate) were excluded. Twenty-Seven (27) States were included as listed in figure 1. All states mandated social distancing universally across their respective territory of the 6 variations listed above on a specific date provided by the IHME data set accessed on June 17, 2020 and maintained them through their initial peak infection rate.

**Figure 1:**
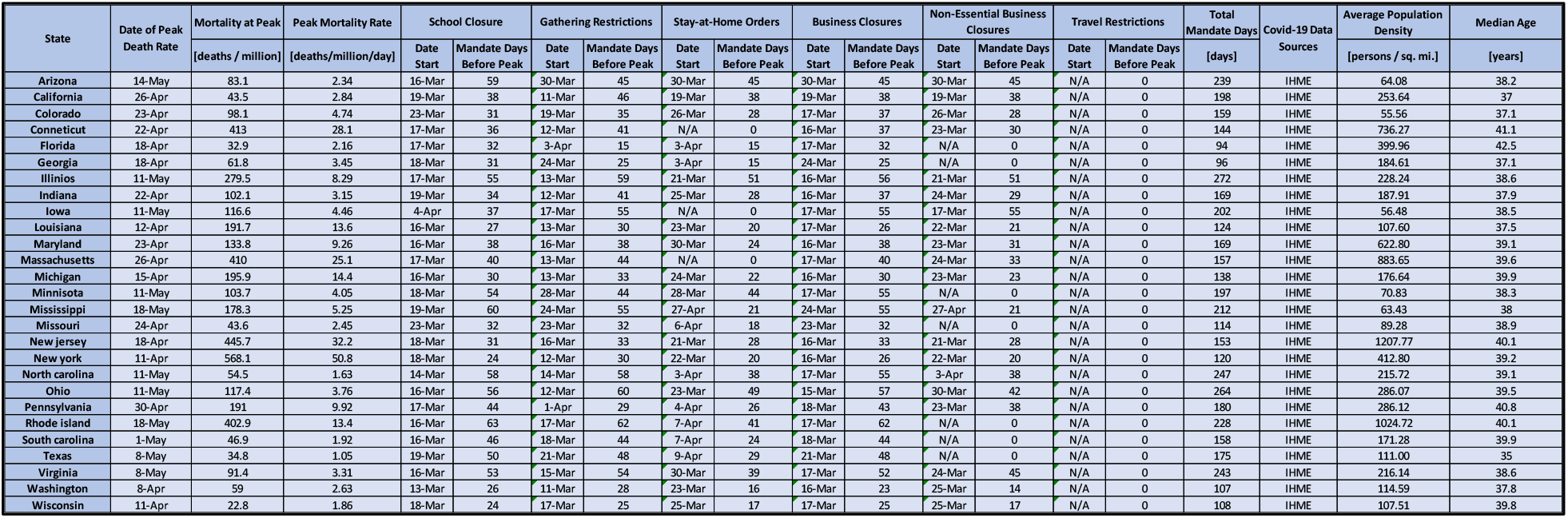
Table of population density, median age, social mandate, and Covid-19 mortality data on 27 states of the USA used in this analysis. Fields with”N/A” indicate the state did not implement that particular form of social mandate.

**Figure 1** captures population density, social mandate form and duration, and Covid-19 mortality and maximum mortality rate data on the 27 states of the USA included in the study. Covid-19 data was obtained from IHME.7 State median age data is obtained from StatsAmerica’s USA States in Profile website for median age in 2019.9 State population density was obtained from the United States Census Bureau 2010 summary.10

The defined endpoints for the analysis included the date of the initial peak mortality in deaths per day for each state. A state’s peak mortality was defined as the highest recorded daily deaths over a seven-day moving-average which was followed by a seven-or-more day decline in mortality with no other discernable peaks (using the same criteria) at the time of accessing the data. The maximum daily mortality rate was used as an easily defined universal milestone in any infectious disease progression to examine the total viral mortality up to the maximum mortality rate.

Additionally, the maximum slope of the Covid-19 mortality was determined by evaluating the maximum of the derivative of the mortality curve. Specifically, this slope was defined as the total recorded mortality five days after the peak-mortality-rate minus the total recorded mortality five days before the peak-mortality-rate divided by the 10-day interval. Both the mortality-at-peak and peak-mortality-rate were normalized by dividing by the population of the selected state.

***Equation 1:*** *Formula to calculate the estimate for the peak-mortality-rate*.

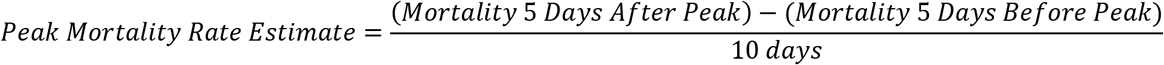

All data was analyzed with Matlab and Microsoft Excel. ^11^ General linear mixed effects (GLME) was used to examine combined effects of the multiple variables. Specifically, Microsoft Excel was used for the bivariate linear regressions and ANOVA summaries for both mortality-at-peak and peak-mortality-rate while Matlab was used to generate the 95% confidence interval bounds on those linear regressions as well as the separate multivariate GLME analyses.

The clinical study was conducted within the ethical principles contained in Declaration of Helsinki, Code of Federal Regulations (CRF), Obligations of Clinical Investigators (21 CFR 812). All data was public and anonymous so no IRB was needed.

Similar to the previous study on European countries, the study conducted two high-level bivariate analyses on this data: one for each output of interest – peak-mortality-rate (PMR) and mortality-at-peak (MAP) – against the *total mandate days*. ^8^ The *total mandate days* is defined the same way as it was in the previous study which is the summation from each form of social-mandate of the total number of days prior to the initial peak that form of social-mandate was implemented. ^8^

***Equation 2****: Formula on how Total Mandate Days are calculated from each form of social-mandate and the number of days prior to the initial peak the mandates were implemented for*.

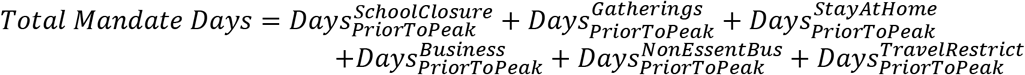

After the bivariate analyses against total mandate days, we then conducted bivariate analyses with the same response variables against population density instead. After the bivariate analyses, two multivariate analyses were conducted. These multivariate analyses used the same response variables as in the previous bivariate studies.

***Equation 3:*** *GLME model for peak-mortality-rate studied in the multivariate analysis*.

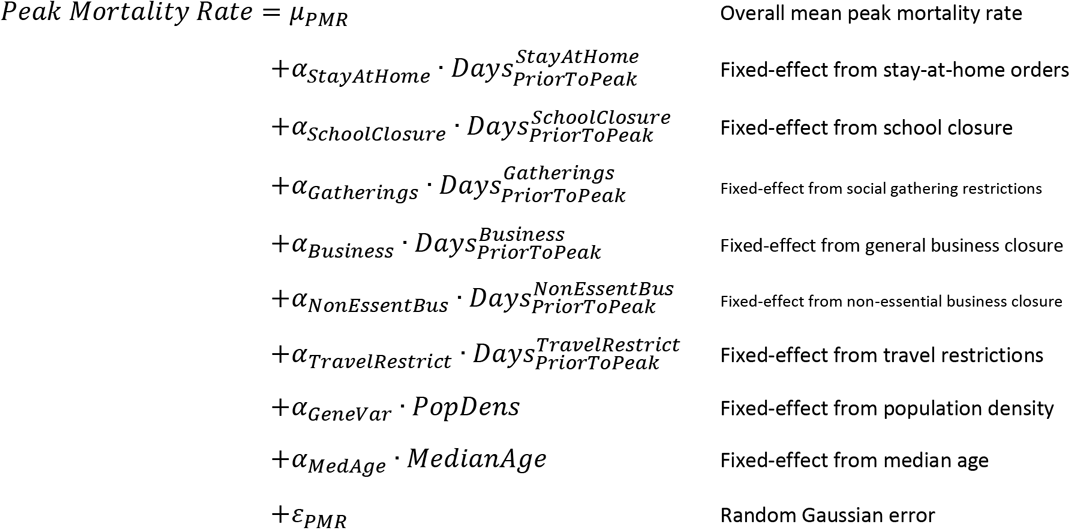

***Equation 4:*** *GLME model for mortality-at-peak studied in the multivariate analysis*.

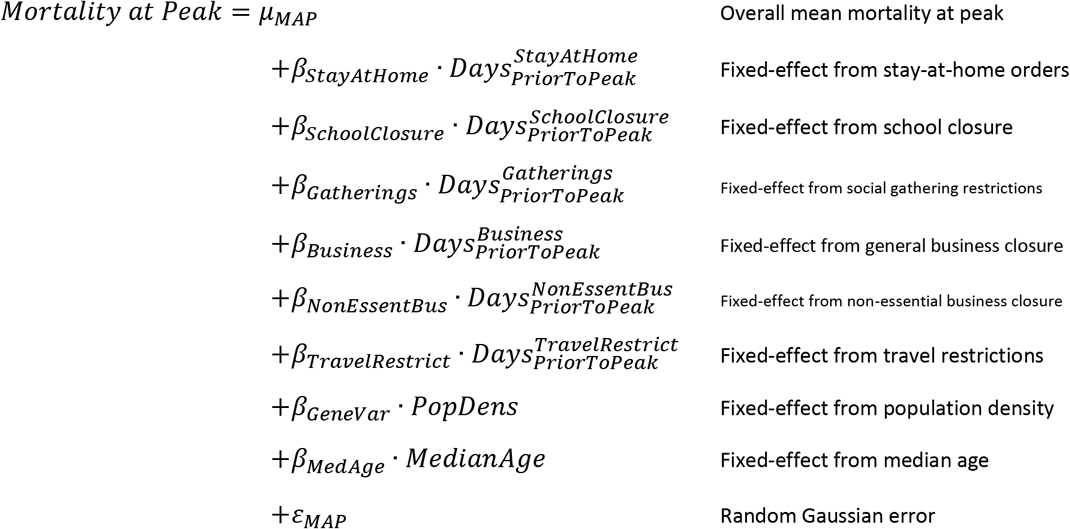

These models were then regressed onto the data from **Figure 1**. Note that where certain social distancing mandates were not implemented, the”Days Prior to Peak” values were coded as zero in modeling to avoid singular data matrices during the regression analysis. The travel restrictions mandate was removed from the analysis since no states implemented strict travel restrictions in the time of this study according to **Figure 1**.

## Results

### Results from Bivariate Analysis, MAP vs Total Mandate Days

Twenty-Seven states were selected June 17, 2020 to examine their collective correlation between standardized mortality and total mandate-days of state directed social distancing directives. All states maintained social distancing directives through the study endpoint.^7^ States were found to have a significant diversity in total mandated intervention over time (mean = 171 total-mandate-days, std-dev = 50 total-mandate-days). The total population studied was 292 million. All states had statistically similar age distributions (mean = 38.6 years-old, std-dev = 1.5 years-old). The population density varied significantly (mean = 304 Pop./square mile, std-dev = 306 Pop./square mile)

Results for the bivariate analysis of mortality-at-peak against total mandate days is captured in **Figure 2**. The correlation of the standardized mortality-at-peak with total-mandate-days of social-distancing mandates prior to the peak was found to be statistically insignificant (R^2^ = 2E-06 with p-val = 0.9946).

**Figure 2:**
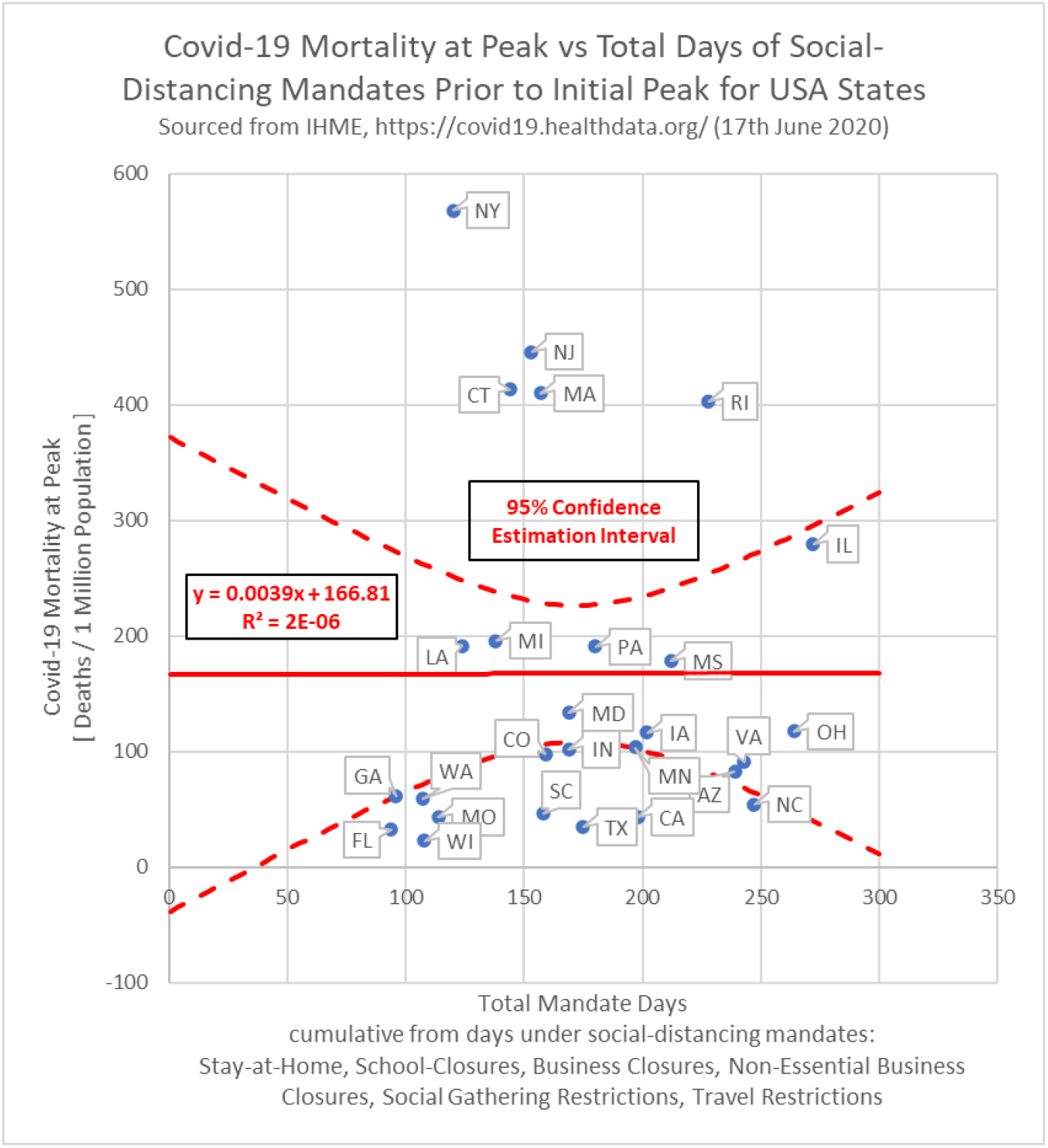
Standardized Covid-19 mortality on the day of peak-mortality-rate correlated to days under state-mandated social distancing directives prior to the peak.

### Results from Bivariate Analysis, PMR vs Total Mandate Days

Results for the bivariate analysis of peak-mortality-rate against total mandate days is captured in **Figure 3**. The correlation of the standardized peak-mortality-rate with total-mandate-days of social-distancing mandates prior to the peak was found to be statistically insignificant (R^2^ = 0.0534 with p-val = 0.2463).

**Figure 3:**
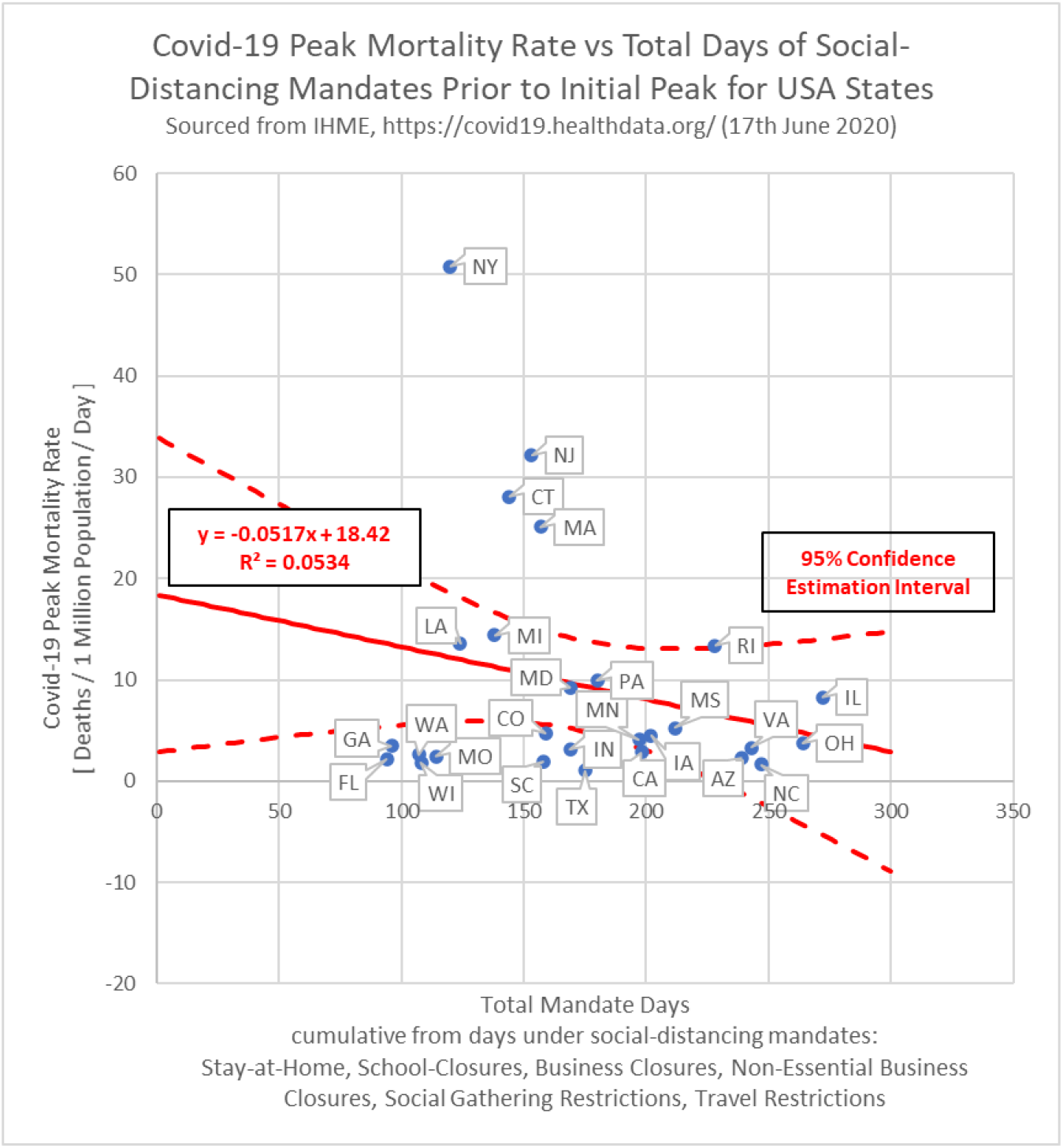
Standardized Covid-19 peak-mortality-rate correlated to days under state-mandated social distancing directives prior to the peak.

### Results from Bivariate Analysis, MAP vs Population Density

Results for the bivariate analysis of mortality-at-peak against average population density is captured in **Figure**. The correlation of the mortality-at-peak with average population density was found to be statistically significant (R^2^ = 0.5238 with p-val = 1.99E-05).

**Figure 4:**
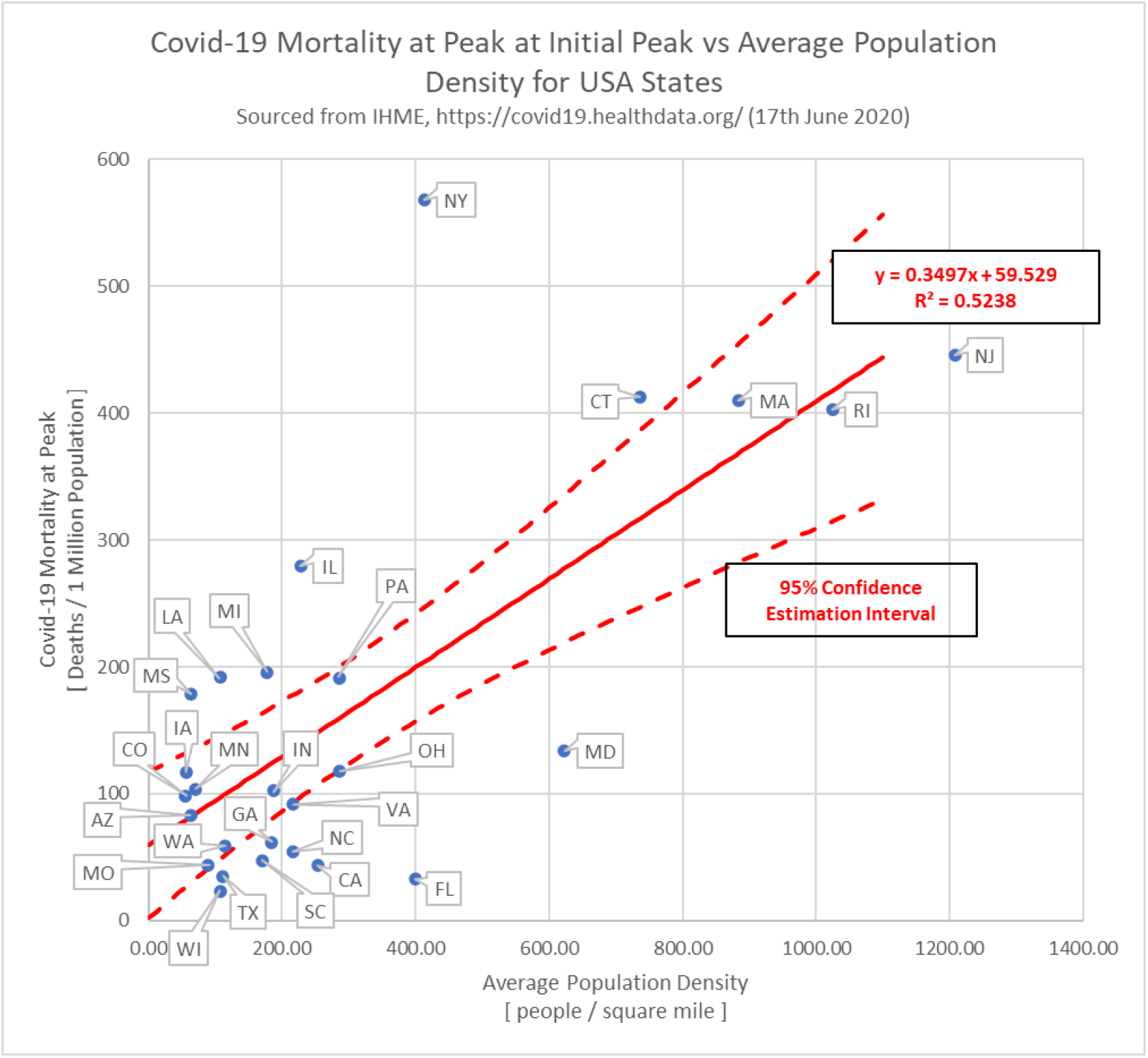
Standardized Covid-19 mortality on the day of peak-mortality-rate correlated to state average

### Results from Bivariate Analysis, PMR vs Population Density

Results for the bivariate analysis of peak-mortality-rate against average population density is captured in **Figure 4**. The correlation of the standardized peak-mortality-rate with total-mandate-days of social-distancing mandates prior to the peak was found to be statistically significant (R^2^ = 0.3814 with p-val = 0.0006).

**Figure 4:**
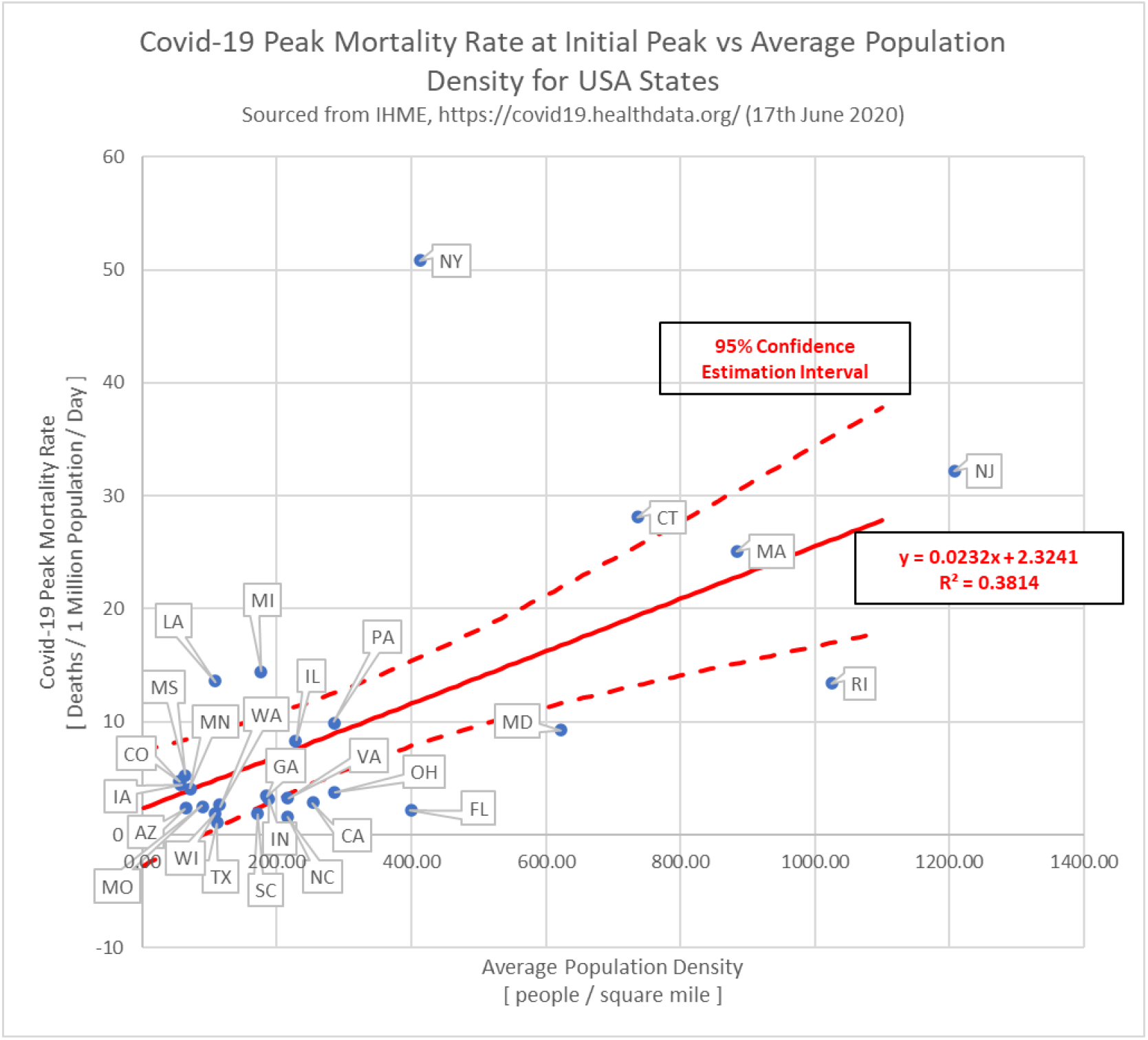
Standardized Covid-19 peak-mortality-rate correlated to state average population density

### Results from Multivariate Analysis for MAP

Results for the multivariate analysis of mortality-at-peak on the date of initial peak-mortality-rate are captured in **Figure 5**. In this study, only one modeled effect was found to be statistically significant at the 5% significance level; this was a state’s average population density (p-val = 0.0004).

**Figure 5:**
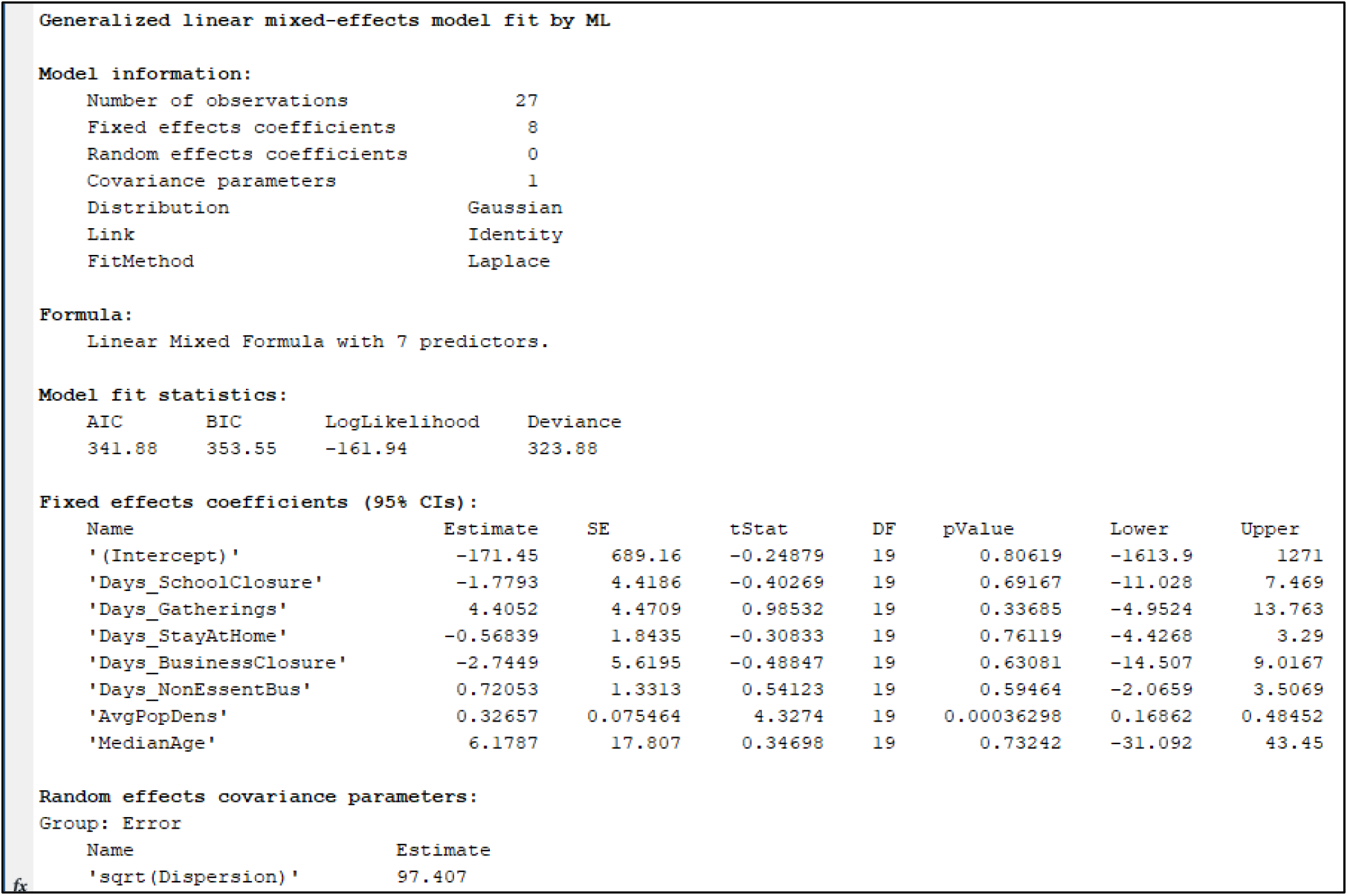
Matlab console output from GLME regression observing mortality-at-peak as the response variable using the mathematical model from Equation 4.

### Results from Multivariate Analysis for PMR

Results for the multivariate analysis of peak-mortality-rate are captured in **Figure 6**. In this study, only one modeled effect was found to be statistically significant at the 5% significance level; this was a state’s average population density (p-val = 0.0025).

**Figure 6:**
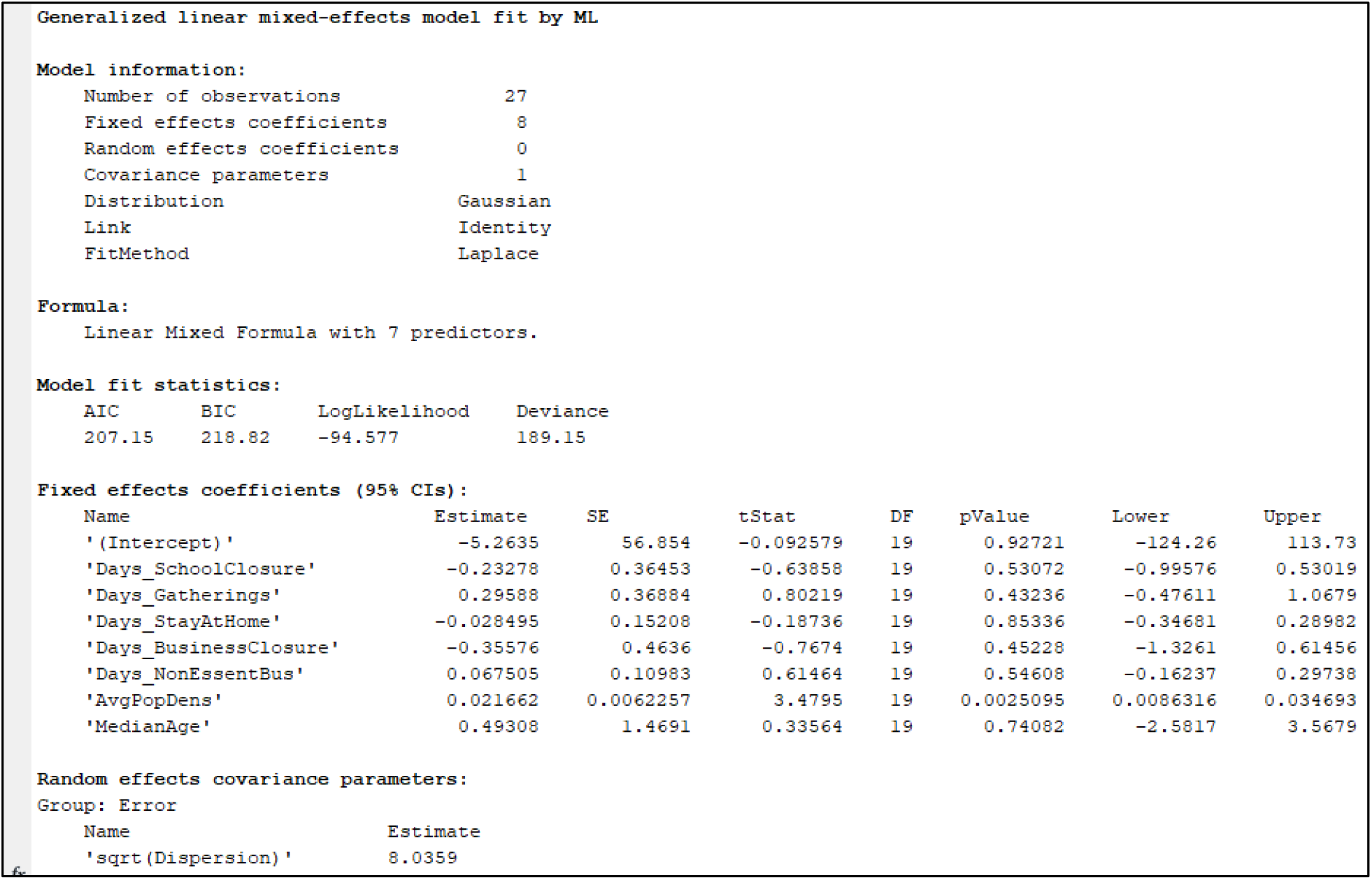
Matlab console output from GLME regression observing standardized peak-mortality-rate as the response variable using the mathematical model from Equation 3.

## Discussion

The analysis appears to show no statistically significant reduction in neither the slope of the Covid-19 mortality rate (p-val = 0.995) nor in Covid-19 mortality (p-val = 0.246) to its defined initial infection peak. There was a significant correlation to both the overall mortality and the maximum mortality rate to state population density. The correlation to population density suggests that the proximity and frequency of social interactions directly affect the mortality rate and overall mortality. However, the evaluation of state mandates suggests that they were unable to reduce the frequency of social interactions to be effective. The results corroborate the findings of the previous study conducted using the same methodology with the IHME data for 12 European countries. ^8^

One of the advantages in this analytical approach is in the design of the social-mandate input variables to be more robust to possible biases caused by different states being in different epidemiologic infectious stages of Covid-19. It is worth noting that quite a few of the states involved in this study have since experience a second wave of infections not included in this analysis. This may be a successive superposition of infectious cycles within the state in sequestered populations or an effect of behavioral changes within the population. As of the drafting of the manuscript on July 19, 2020, Six States; Arizona, California, Florida, Mississippi, South Carolina, Texas have had secondary peaks of maximum infection rates which eclipsed the primary peak used in this analysis. Only their initial peaks were the subject of this study and they were not revised to the new peak nor excluded. Mandates and social behaviors changed substantially during the interim between the first maximum and could confound the analysis. Furthermore, the strong correlation to population density demonstrating a likely effect of personal interaction frequency which assists in validating the methodology. It is also worth noting that start dates of social-mandate state implementation does not necessarily imply a certain degree of compliance of the population with enacted mandates.

## Conclusions

How early before the initial peak an investigated form of social-mandate was implemented by a USA state was found to be statistically insignificant in its effects on peak-mortality-rate and mortality-at-peak in both the bivariate studies and the multivariate studies. (The lowest associated p-value with social-mandate terms across all studies above was p-val > 0.2463 in the bivariate study of studying standardized Covid-19 peak-mortality-rate against total-mandate-days.) However, the average population density was found to be a statistically significant factor for both peak-mortality-rate and mortality-at-peak of a state in both the bivariate and multivariate studies. (The largest associated p-value with average population density across all studies above was p-val = 0.0025 in the multivariate study with peak-mortality-rate.)

What we could conclude from this study is that the timing of social-mandates alone is not enough to explain the variability in Covid-19 peak-mortality-rate and mortality-at-peak numbers between states. However, a state’s average population density is a significant factor and should be accounted for in one way or another in current and future investigations to explain the variability in Covid-19 peak-mortality-rate and mortality-at-peak numbers between states.

Factors which influence the effectiveness of social distancing interventions include the reproduction number (R_0_) of the virus, the mortality rate, and the mandate effectiveness of the isolation.^12,13^ A reduction of the slope of the maximum infection rate without a change in mortality is consistent with theory of viral infectious epidemiology.^14^ Reductions in total mortality due to reduced healthcare system or advancements in medical care did not materialize in this analysis. However, the time from the probable first infection to the peak of the infection rate was likely only around 3 months. It is possible that if the viral mortality estimated at 0.6% to 1.2% or the viral reproduction R_0_, estimated between 2.8 and 3.3, were significantly higher may have produced a correlation.^11,12^ Mandated interventions which were significantly more isolating may have produced a correlation to the mortality.

## Data Availability

all data is publically available http://www.healthdata.org/covid

List of Abreviations:

- FDA

- Food and Drug Administration
- CFR

- Code of Federal Regulations
- IHME

- Institute of Health Metrics and Evaluation
- GLME

- General Linear Mixed-Effects
- PMR

- Peak-mortality-rate due to Covid-19, standardized by state total population, as calculated from **Equation 1** using data from IHME
- MAP

- Mortality-at-peak due to Covid-19, standardized by state total population as reported in referenced IHME data

## Declarations

### Ethics approval and Consent to participate

- This clinical study was conducted in accordance with the ethical principles contained within Declaration of Helsinki, Protection of Human Volunteers (21 CFR 50), Institutional Review Boards (21 CFR 56), and Obligations of Clinical Investigators (21 CFR 812). No IRB was required with the use of depersonalized public data.

### Consent for publication

- Not Applicable. Depersonalized data made available for download on IHME Websites can be used, shared, modified or built upon by non-commercial users via the Creative Commons Attribution-NonCommercial 4.0 International License. https://creativecommons.org/licenses/by-nc/4.0/

### Availability of data and materials

- Depersonalized data made available for download on IHME Websites can be used, shared, modified or built upon by non-commercial users via the Creative Commons Attribution-NonCommercial 4.0 International License. https://creativecommons.org/licenses/by-nc/4.0/

### Authors’ Contributions

- Sean McCafferty – Design, drafting, analysis, and interpretation
- Sean Ashley – Drafting, analysis, and interpretation

Acknowledgements
- None.

